# The Correlation Between Brain Performance Capacity and COVID-19: A Cross-sectional Survey and Canonical Correlation Analysis

**DOI:** 10.1101/2022.01.29.22270064

**Authors:** Y Liu, X Chen, JS Xian, R Wang, K Ma, K Xu, X Yang, FL Wang, N Mu, S Wang, Y Lai, T Li, CY Yang, YL Quan, H Feng, LH Wang, TN Chen

## Abstract

**Objective:** To generate a concept of brain performance capacity (BPC) with sleep, fatigue and mental workload as evaluation indicators and to analyze the correlation between BPC and the impact of COVID-19.

**Methods:** A cluster sampling method was adopted to randomly select 259 civil air crew members. The measurements of sleep quality, fatigue and mental workload (MWL) were assessed using the Pittsburgh Sleep Quality Index (PSQI), Multidimensional Fatigue Inventory (MFI-20) and NASA Task Load Index. The impact of COVID-19 included 7 dimensions scored on a Likert scale. Canonical correlation analysis (CCA) was conducted to examine the relationship between BPC and COVID-19.

**Results:** A total of 259 air crew members participated in the survey. Participants’ average PSQI score was 7.826 (SD = 3.796), with 49.8% reporting incidents of insomnia, mostly of a minor degree. Participants’ MFI was an average 56.112 (SD = 10.040), with 100% reporting some incidence of fatigue, mainly severe. The weighted mental workload (MWL) score was an average of 43.084 (SD = 17.543), with reports of mostly a mid-level degree. There was a significant relationship between BPC and COVID-19, with a canonical correlation coefficient of 0.507 (*P*=0.000), an eigenvalue of 0.364 and a contribution rate of 69.1%. All components of the BPC variable set: PSQI, MFI and MWL contributed greatly to BPC, with absolute canonical loadings of 0.790, 0.606 and 0.667, respectively; the same was true for the COVID-19 variable set, with absolute canonical loadings ranging from 0.608 to 0.951.

**Conclusion:** Multiple indicators to measure BPC and the interrelationship of BPC and COVID-19 should be used in future research to gain a comprehensive understanding of anti-epidemic measures to ensure victory in the battle against the spread of the disease.

## INTRODUCTION

Brain performance capacity, a complex concept, is associated with memory, cognitive capacities, attention, and work efficiency. This has been tested by research and resulted in much evidence. As early as 1975, Kahn, R L(1) held the idea that impaired memory and depression were the products of poor brain performance. L. Nyberg also argued that brain maintenance constituted the primary determinant of successful memory aging (2). In modern society, it is vital for optimal performance to enhance or preserve the cognitive performance of personnel working in stressful, demanding and/or high-tempo environments (3). Furthermore, the ability to focus attention on tasks may contribute to peak brain performance and high-level work efficiency. A. Yamashita found that individuals with attention-deficit/hyperactivity disorder (ADHD) spent more time and energy maintaining optimal and effective performance behavior during tasks that required sustained attention (4).

It is well known that in some professional contexts, such as aviation, where staying in good physical condition is requisite, there are additional traits, such as BPC, that are important factors for success. However, how can this aspect be valuated? We have a remarkably poor understanding of what the direct indicator might be in measuring BPC.

However, there are some factors, such as sleep, fatigue-free state, and appropriate level of mental workload, which are thought to be related to performance; vigilant attention and long memory are also traits that may indirectly boost peak brain performance. Much evidence suggests that the right number of hours and quality of sleep improves working memory (5), vigilant attention (6), concentration (7), but fatigue and high-level mental workload impair them (8-13). Taking flight crews as an example, members and particularly pilots, are submitted to high, exigent work demands and must manage multiple tasks at the same time. In addition, they are continuously exposed to stimuli that compete for their attention and ability to manage their resources to make the right decisions. These task-oriented responsibilities are further complicated by complex flying factors, such as long trips and varying shift flights.

It was reported that the majority of accidents, 60%∼90%, are attributed to “human error” (14-16) and occur when flight crews are subjected to a high and intensive mental workload level (17). Thus, BPC is a prerequisite to ensure flight safety. The COVID-19 pandemic has had a huge impact on people’s lives, the economy, physical-mental health, and BPC, since the outbreak began in 2020. Because of the instability and resurgence of the disease, it will continue to have a negative impact for some time even though we have entered a new period known as the normalized era of COVID-19. Like the general population, aviation crews have been exposed to high-risk environments COVID-19 environments for a long time. Thus, this study aimed to assess the impact in the following ways: 1) update information about the sleep quality, fatigue and mental workload of commercial flight crews in the normalized era of COVID-19 in China, and 2) generate a dependent variable cluster named BPC with indicators such as sleep quality, fatigue and mental workload and to explore the relationship between BPC and the impact of COVID-19 in this new era.

## METHODS

### 2.1 Participants and study design

This was a cluster sampling and cross-sectional study. The recommended minimum sample size was 200, which was 10 times the number of MFI-20 for a maximum number of entries in this study (18). Estimating a 20% invalid survey response rate, the expected sample was at least 240. The final sample was 259.

*Questionnaire Star* was used to collect sociodemographic data and key targets (sleep, fatigue and mental workload) from 25 November to 2 December 2021. Inclusion criteria: a. volunteer to participate in this survey, b. have the ability to use basic modern information technology, such as WeChat or computers. Exclusion criteria: a. not full-attended during the investigation request period due to any reasons, b. have physical and mental illness. The eligible participants were asked to answer all the questions about sleep, fatigue and mental workload in the previous month. Subjects logged on to Questionnaire Star via WeChat with a smartphone or computer. Individuals completing the survey received a red envelope with a random amount of money as compensation from *Questionnaire Star*.

Demographics including gender, age, BMI, position, and social jet lag, were collected. Respondents were also asked about the severity of the impact of COVID-19 on them and scored their responses on a Likert scale (1= never, 5 = extremely serious). This scale included seven dimensions: fear of self-infection (C1); fear of ineffective prevention and control (C2); physical unwellness caused by regular nucleic acid testing (C3); increased workload of personal protective equipment (C4); sleep quality (C5); mental fatigue (C6); and physical fatigue (C7).

### 2.2 Scales to measure indicators of BPC

#### 2.2.1 Fatigue Assessment

A multidimensional fatigue inventory (MFI-20) was conducted to evaluate the fatigue of commercial pilots. The scale was created by Ellen Smets and colleagues (19) in 1995 and has been translated into various languages by other researchers worldwide. The 20-item scale was designed to evaluate five dimensions of fatigue: general fatigue, physical fatigue, reduced activity, reduced motivation and mental fatigue. Twenty questions were rated on a 5-point scale (“yes, that is true” to “no, that is not true”). The higher the total score was, the greater the fatigue. The total MFI scores range from 0 to 100 and are interpreted as no fatigue (0-20), minor fatigue (21-39), serious fatigue (40-59), severe fatigue (60-79) and extremely severe fatigue (80-100). This scale has been verified by Chinese scholars and can be used to test the fatigue of Chinese pilots (20).

#### 2.2.2 Sleep Assessment

The Pittsburgh Sleep Quality Index (PSQI) was used to assess the sleep quality of the participants. This mature scale, widely used to evaluate sleep quality, has 7 items: sleep latency, sleep duration, sleep quality, habitual efficiency, sleep disturbance, use of sleeping medications and daytime dysfunction^21^. Total PSQI scores range from 0 to 21, and 8 points or more is regarded as a sleep disorder, 8-10 points as a minor sleep disorder, 11-15 points as a serious sleep disorder, and 16 points or more as a severe sleep disorder (22). The higher the score was, the worse the sleep quality.

#### 2.2.3 Mental Workload Assessment

NASA Task Load Index (NASA TLX) (23) was utilized to measure the mental workload of the participants. It is a tool for measuring and conducting a subjective mental workload (MWL) assessment. It allows the subject to determine the MWL of a participant while they are performing a task. It rates performance across six dimensions to determine an overall workload rating. The six dimensions are as follows:

1. Mental demand: How much thinking, deciding, or calculating was required to perform the task?
2. Physical demand: How much physical activity and intensity was required to complete the task?
3. Temporal demand: What kind of time pressure was involved in completing the task?
4. Effort: How hard does the participant have to work to maintain his or her level of performance?
5. Performance: How successful was the individual in completing the task?
6. Frustration level: How insecure and discouraged or secure and content did the participant feel during the task?

Each subscale is presented to the participants either during or after the experimental trial. They are asked to rate their score on an interval scale ranging from low (1) to high (100). The TLX also employs a paired comparisons procedure. This involves presenting 15 pairwise combinations to the participants and asking them to select the scale from each pair that has the most effect on the workload during the task under analysis. This procedure accounts for two potential sources of between-rater variability: differences in workload definition between the raters and differences in the sources or workload between the tasks. The total MWL score is 0-100 points. The higher the scores, the higher the mental workload. In this study, we used the weighted workload score. This scale has been verified by Chinese scholars and can be used to test the mental workload of Chinese pilots.

### 2.3 Statistical analyses

The statistical analyses were carried out by IBM SPSS Statistics 26.0. All answer sheets were error-checked by the study team. The QQ chart and Bonferroni test were used to check the former’s normality and homogeneity of the variance. Canonical Correlation Analysis (CCA), a multivariate analysis of the association between sets of multiple independent and dependent variables, was administered to explore the relationship between BPC and the impact of COVID-19. Each set can be named. The canonical correlation coefficient (Rc) maximizes the correlation between the canonical variate obtained by synthesizing multiple independent and dependent variables. In this study, we constructed an independent variable set named *COVID-19* that affected the dependent variable set: sleep quality, fatigue state and mental workload, which was named *brain performance capacity (BPC)*. The relationships between the 2 sets of variables were interpreted using canonical loadings to determine how much each variable contributed to its own set and using canonical cross-loadings to determine how much each variable contributed to the other set. The significance level (p) of the correlation was set at 0.05, and a loading >0.30 was regarded as an important contribution.

## RESULTS

### 3.1 Participant

In total, 259 flight crew members participated, and the number of valid responses was 100% without any missing data. Overall, 51.7% of the flight crew members (n = 134) were male, and 48.3% were female (n=125). The age of the participants ranged from 17 to 55 years, with an average of 28.204 ± 6.728. The weighted NASA-TLX scores ranged from 0 to 100, and the mean score was 43.084 ± 17.543. The PSQI scores ranged from 0∼22 with a mean score of 7.826 ± 3.796, and the MFI score ranged from 22 to 77 with a mean score of 56.112 ± 10.040. The insomnia incidence of the participants was 49.8%, mainly minor insomnia (30.2%), followed by serious (15.8%) and severe insomnia (1.9%), with a significant difference among the insomnia degrees (X^2^=134.745, P=0.000). The fatigue incidence was 100%, mainly severe fatigue (47.9%), followed by serious (42.1%) and minor fatigue (10%), with significant difference among them (X^2^=64.548, P=0.000). The distribution of civil aviation crew BPCs is detailed in Table 1.

**Table 1.** Distribution of Civil Aviation Crews’ BPC (n=259)

| Item |  | PSQI<br>(Mean ± SD) | MFI<br>(Mean ± SD) | MWL<br>(Mean ± SD) |
| --- | --- | --- | --- | --- |
| **gender** | Male(n=134) | 8.08±4.04 | 53.99±11.04 | 44.06±18.65 |
|  | Female(n=125) | 7.55±3.50 | 58.39±8.30 | 42.04±16.28 |
|  | t value | 1.13 | -3.65 | 0.93 |
|  | P value | 0.26 | 0.00 | 0.35 |
| **age/y** | 28.20±2.73 | 20.38±7.35 | -27.91±12.3 | -14.8±18.75 |
|  | t value | 44.63 | 36.52 | -12.77 |
|  | P value | 0.00 | 0.00 | 0.00 |
| **BMI** | 21.11±3.89 | 13.28±5.42 | -35±11.17 | -21.9±17.93 |
|  | t value | 39.42 | -50.43 | -19.73 |
|  | P value | 0.00 | 0.00 | 0.00 |
| **position** | pilots(n=78) | 7.60±4.08 | 54.06±11.7 | 44.18±18.18 |
|  | flight attendant(n=138) | 7.96±3.87 | 56.96±9.1 | 42.88±16.73 |
|  | security(n=43) | 7.79±3.00 | 57.09±9.31 | 41.75±19.17 |
|  | F value | 0.23 | 2.35 | 0.28 |
|  | P value | 0.80 | 0.10 | 0.75 |
| **driving experience/y** | ≥5(n=187) | 7.58±3.41 | 56.33±10.22 | 42.29±17.32 |
|  | >5(n=72) | 8.47±4.60 | 55.56±9.61 | 45.14±18.06 |
|  | t value | -1.50 | 0.56 | -1.17 |
|  | P value | 0.14 | 0.58 | 0.24 |

### 3.2 CCA of BPC and COVID-19

#### 3.2.1 Spearman’s Correlation

Table 2 shows the correlation coefficient matrix among the three components of BPC scales and the seven domains of COVID-19 impact on the civil aviation crews. Except for C4 and MFI, the correlation coefficients among the components were significant, from the lowest absolute value of 0.140 (*P*<0.05) to the largest absolute value of 0.896 (*P* < 0.01).

**Table 2.**
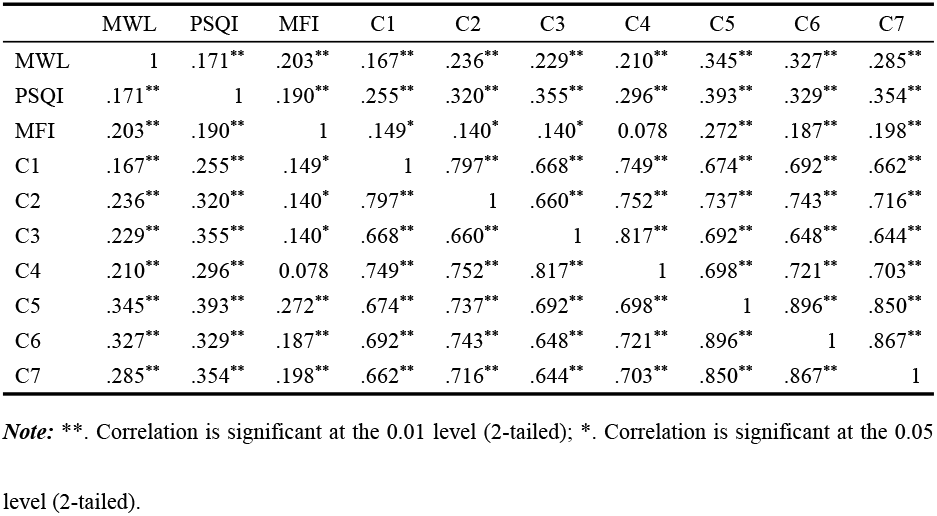
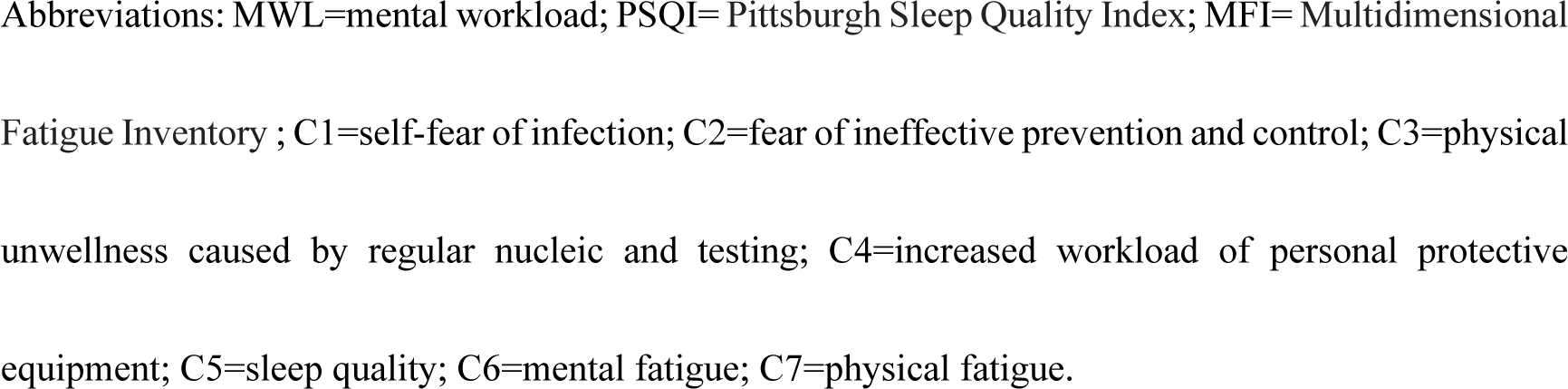
Spearman’s Correlation between BPC and COVID-19

#### 3.2.2 Canonical Correlation

The CCA was performed between BPC variables and COVID-19 variables, and the outcome contained 3 pairs of canonical variables. The correlation coefficient of the first pair of canonical variates was statistically significant (*P*=0.000 < 0.05), with the correlation coefficient having a value of 0.517, an eigenvalue of 0.364, and a contribution rate of 69.1%. Thus, only the first pair of canonical variables was analyzed further (Table 3).

**Table 3.** Results of CCA between BPC and COVID-19

|  | Correlation | Eigenvalue | Wilks<br>Statistic | *F* | *u* | *P* |
| --- | --- | --- | --- | --- | --- | --- |
| 1 | 0.517 | 0.364 | 0.691 | 4.681 | 21.000 | 0.000 |
| 2 | 0.206 | 0.044 | 0.943 | 1.247 | 12.000 | 0.247 |
| 3 | 0.124 | 0.016 | 0.985 | 0.789 | 5.000 | 0.558 |

According to Table 4, which shows the standardized canonical correlation coefficients, a set of linear combinations within a group of typical variables can be obtained:

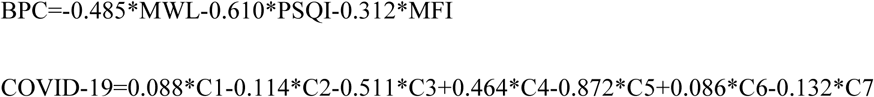

The proportion variance explained by the BPC set was 48.3%, and that explained by the COVID-19 set was 58%.

**Table 4.** Standardized Canonical Correlation Coefficients of BPC set and COVID-19 set

| Variable set | Variable | Standardized Canonical<br>Correlation Coefficients | Proportion Variance<br>of Explained |
| --- | --- | --- | --- |
| BPC | MWL | -0.485 | 0.483 |
|  | PSQI | -0.610 |  |
|  | MFI | -0.312 |  |
| COVID-19 | C1 | 0.088 | 0.580 |
|  | C2 | -0.114 |  |
|  | C3 | -0.511 |  |
|  | C4 | 0.464 |  |
|  | C5 | -0.872 |  |
|  | C6 | 0.086 |  |
|  | C7 | -0.132 |  |

Abbreviations: BPC=brain performance capacity; MWL=mental workload; PSQI= Pittsburgh Sleep Quality Index; MFI= Multidimensional Fatigue Inventory; C1=self-fear of infection; C2=fear of ineffective prevention and control; C3=physical unwellness caused by regular nucleic and testing; C4=increased workload of personal protective equipment; C5=sleep quality; C6=mental fatigue;
C7=physical fatigue.

Fig 1 shows the canonical loadings of the BPC set and the COVID-19 set. All MWL, PSQI and MFI contribute greatly to BPC, with an absolute value of the lowest canonical loading coefficient of 0.606. All components of COVID-19 contribute greatly to COVID-19, with absolute values of canonical loading coefficients from 0.608 to 0.951. This indicates that the indicators MWL, PSQI and MFI could well inflect the BPC. Similarly, C1, C2, C3, C4, C5, C6 and C7 present the impacts of COVID-19 on civil aviation crews.

**Figure 1:**
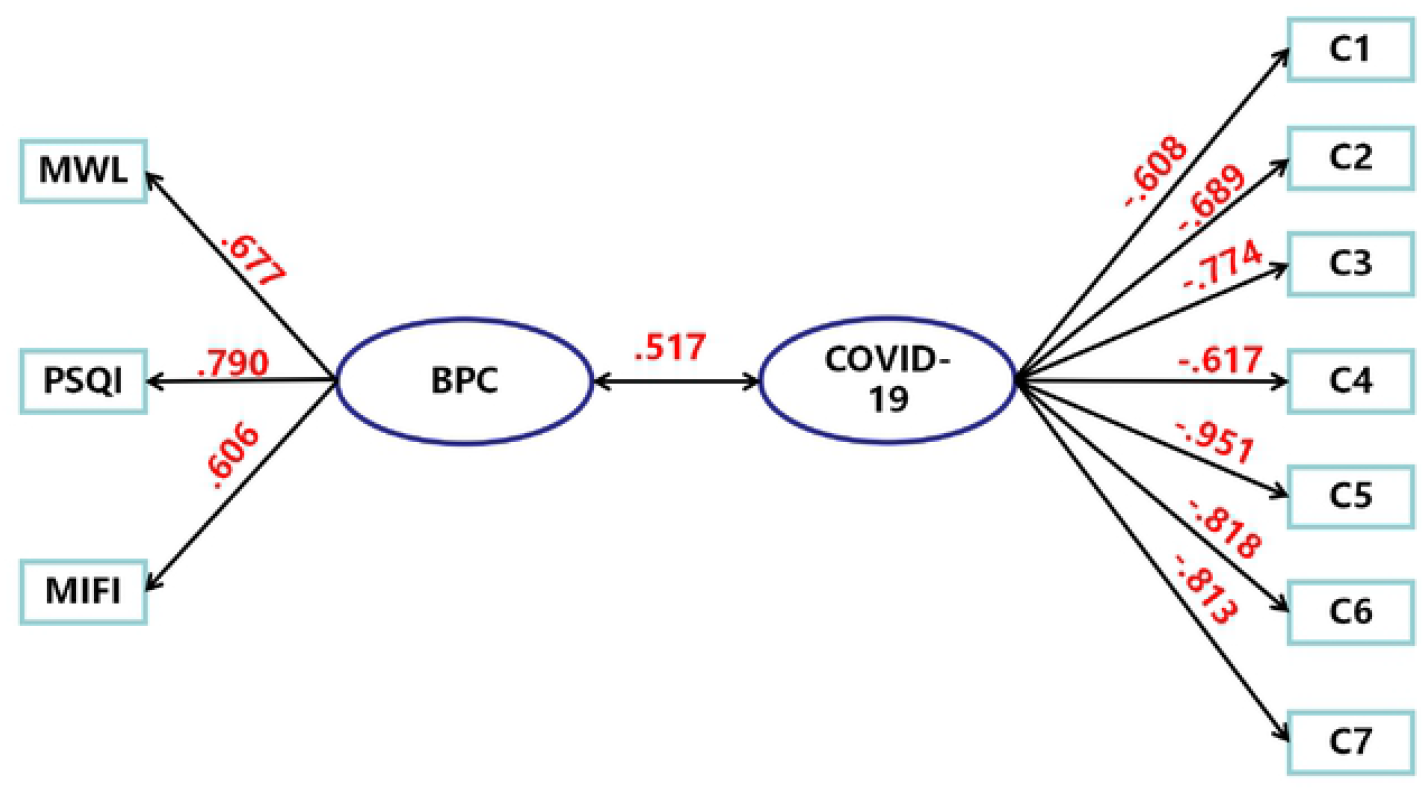
CCA of BPC and COVID-19 Abbreviations: CCA=canonical correlation analysis; BPC=brain performance capacity; MWL=mental workload; PSQI= Pittsburgh Sleep Quality Index; MFI= Multidimensional Fatigue Inventory; C1=self-fear of infection; C2=fear of ineffective prevention and control; C3=physical unwellness caused by regular nucleic acid testing; C4=increased workload of personal protective equipment; C5=sleep quality; C6=mental fatigue; C7=physical fatigue.

## DISCUSSION

This study updated the data on civil aviation crews regarding sleep quality, fatigue and mental workload in the new stage of normalized epidemic era of COVID-19 in China. The insomnia incidence was 49.8%, and gender, position and driving experience had no significant impact. There was a high level of fatigue, with a 100% incidence reported by participants, which was much higher than that reported by US army aviators (24). Men were more tired than women. The MWL was reported to be mid-level, with a weighted score of 43.084 ± 17.543. Furthermore, the older the age and the higher the BMI were, the worse the sleep, the more fatigued and the higher the mental workload level.

It also first reported here that a comprehensive evaluation of BPC can be made through the assessment index cluster we built. Many studies have reported the effects of sleep quality, fatigue and mental workload on individuals’ memory, attention, reaction, and mental health, which could directly or indirectly respond to brain performance. *Yamashita, A* hold the idea (25) that individuals with attention-deficit/hyperactivity disorder (ADHD) have unstable and unreliable behaviors related to the state of their brains, whereas those without ADHD have an optimal brain state that guides their more stable and reliable behavior. The brain has plasticity. L. L. Beason-Held (26) tested the hypotheses that stable task performance could be supported by stable brain performance or that stable performance could be maintained by changing brain functions if the change was a compensatory reorganization of function. We also found that sleep quality, fatigue and mental workload responded well to BPC. Each indicator contributed greatly to BPC, as there were strong loadings of MWL (loading=-0.677), PSQI (loading=-0.790), and MFI (loading=-0.606). As concluded by *Walker, Matthew P*, et al., the brain under sleep deprivation exhibits inattention, decreased working memory, and insufficient motivation to learn (27). We also found that the higher the MWL level was, the lower the BPC; the worse the sleep quality was, the lower the BPC; and the more fatigued an individual was, the lower his or her BPC. These findings may provide new perspectives on how to evaluate BPC.

Additionally, this study highlighted the impact of COVID-19 on civil flight crews. The COVID-19 pandemic has had a profound impact on the aviation industry, leading to job loss, furlough, disruption of careers and training trajectories and uncertainties, even leading to self-harm among airline employees (28). In addition, there is evidence that COVID-19 induces fears, including fear of infection and fear of passing the infection to others due to ineffective prevention and control measures from the government. Due to the vagaries of the epidemic, people receive nucleic acid tests (NATs) on a fixed schedule or and at any time during the period of normalized control of the epidemic in China. Special occupations in the airlines and health care require not only masks but also personal protective equipment such as gloves, gowns and hats, which is bound to increase the workload of these employees. COVID-19 causes sleep problems such as difficulties initiating and maintaining sleep (29) and fatigue, including mental fatigue and physical fatigue (30). With the statistical analysis of canonical loadings of the COVID-19 set, we also found that the fear of infection(C1), fear of ineffective prevention and control(C2), physical unwellness caused by NAT(C3), increased workload of personal protective equipment(C4), sleep quality(C5), mental fatigue(C6) and physical fatigue(C7) all contributed greatly to the impact of COVID-19, with a loading of -0.608, -0.689, -0.774, -0.617, -0.951, -0.818 and -0.813, respectively. The more severe the impact of COVID-19 was, the worse the BPC.

We also explored the relationship between BPC and the impact of COVID-19 with CCA. We found that a moderate positive correlation existed between BPC and the impact of COVID-19, with a correlation of 0.517. The correlation indicated that the more severe the impact of COVID-19 was on civil aviation crews, the stronger the BPC. However, the pandemic has been in a normalized control period in China. As of 24:00 on November 25, 2021, there had been 860 confirmed cases (including 8 severe cases), 93,087 cured and discharged cases, 4,636 deaths, 98,583 confirmed cases and 4 suspected cases; a total of 1,315,485 close contacts had been traced, and 28,342 close contacts were still under medical observation, according to a report from the National Health Commission of the People’s Republic of China (31). COVID-19 was still a predictor of BPC. This finding may suggest that there is a long way to go in the fight against COVID-19, and the negative influences should not be underestimated. There are some valid concerns about how to boost the BPC to respond to the impact of COVID-19 on individuals.

This study has limitations that need to be considered. First, this was a cross-sectional study, so it cannot provide a cause-and-effect relationship. Because BPC is dynamic, more rigorous study designs, such as cohort studies, should be conducted in the future. Second, although sleep, fatigue and mental workload were successfully used as evaluation indicators of BPC, the assessment tools in this study were mainly scales that are subjective. Therefore, more evaluation indicators of BPC should be explored and combined with objective evaluation indicators.

## CONCLUSION

BPC could be assessed by sleep quality, fatigue and mental workload. In the normalized epidemic era of COVID-19 in China, the pandemic can positively affect the level of brain performance of commercial flight crews. Considerations including multiple indicators to measure BPC and the relationship between BPC and COVID-19 should be considered in future research to gain a comprehensive view of anti-epidemic measures as soon as possible.

## Data Availability

All relevant data are within the manuscript and its Supporting Information files.

## Ethical approval and consent to participate

Guided by the 2000 Declaration of Helsinki for ethical standards, the protocol was approved by the Ethics Committee of the First Affiliated Hospital of Army Medical University, PLA (Ref: KY2021047). Informed consent was obtained from the participants prior to their participation. There, they were able to read a 2-page informed consent form. After reading the consent, they could decide whether to take part in this survey. Those who agreed to participate in the study were required to electronically sign the consent form and then to complete the questionnaire; otherwise, the Questionnaire Star would end the interview directly.

## Competing interesting

The authors declare that they have no competing interests. All authors have read the manuscript and agreed to publish.

## Clinical Trial Registry

This study is registered in the Chinese Clinical Trial Registry, number ChiCTR2100053133.

## Authors’ contributions

HF, TNC and LHW conceived, designed, and supervised the study; YL, XC, YL, NM, KX collected the data; YL, JSX, FLW and TL finalized the analysis, designed the study and interpreted the findings; YL and XC drafted the manuscript; YLQ, SW, RW and CYY interpreted the findings and helped revise drafts of the manuscript. all authors have read and approved the final version of the manuscript, and agree with the order of presentation of the authors.

